# Conditional Survival and Nomogram for Elderly Non-Metastatic Colon Cancer Patients Following Colectomy

**DOI:** 10.1101/2024.04.09.24305543

**Authors:** Yadong Gao, Huimin Wang, Yi Zhang, Jing Zhao, Sujuan Feng, Jianwei Qiu

## Abstract

**Background:** This study aimed to evaluate the conditional survival (CS) of elderly patients with non-metastatic colon cancer who underwent colectomy and build conditional nomograms that can accommodate varying survival periods and estimate survival rates.

**Methods:** Data from 9302 patients between 2004 and 2017 were obtained from the Surveillance, Epidemiology, and End Results database. CS was used to assess overall survival and colon-specific survival rates in patients who survived beyond a certain time period. Cox regression was used to select factors for nomogram development, and performance was evaluated using area under the receiver operating characteristic curve (AUC), calibration plot, and decision curve analysis (DCA).

**Results:** The 5-year conditional overall survival rates initially increased slightly but then decreased over time. The rates at different time points after diagnosis (baseline and 1-5 years) were 62.5, 63.2, 62.8, 62.1, 61.6, and 59.8%. In contrast, 5-year conditional colon-specific survival rates consistently improved over the same period. These rates were 78.1, 80.9, 84.2, 86.9, 89.3, and 90.9%, respectively. Nomograms were developed for baseline measurements and for patients who survived 1, 3, and 5 years. The performance of these nomograms, assessed using AUC, calibration curves, and DCA, indicated good predictive capabilities.

**Conclusion:** CS provides valuable information on the medium- and long-term survival probabilities of elderly patients with non-metastatic colon cancer who underwent colectomy. The developed conditional nomograms allowed for the estimation of survival probabilities across different timeframes, facilitating a comprehensive understanding of prognosis and collaborative decision making.

## 1. Introduction

Colon cancer is the third most common and deadly form of cancer worldwide, accounting for 10% of all cancer cases^1^. It mainly affects the elderly population, with most patients aged > 60 years and a significant number aged > 75^2^. As the global population ages, a significant increase in the prevalence of colon cancer is expected among older individuals. In the United States alone, the elderly population (65 years and older) is expected to increase by 20% by 2030, leading to a parallel surge in colon cancer cases^3^. Additionally, the number of adults aged ≥ 85 years is expected to triple to 19 million by 2060, further contributing to the rise in colon cancer cases. While there is no standardized definition of “elderly” in relation to colon cancer, it generally includes individuals aged 65-75 or older^4–6^.

Surgery is the preferred treatment for stage I–III colon cancer. Advances in medical standards have made radical surgery more feasible in elderly patients. Studies have shown an increase in the rate of radical surgery among colon cancer patients aged ≥ 75 years who were diagnosed with stage I or II^7^. Surgical resection has been found to improve the prognosis of elderly colon cancer^8–9^. Improved perioperative management and comprehensive geriatric assessments have contributed to better tolerance to surgical treatment and improved quality of life in the elderly^10^. These factors have led to prolonged survival of elderly patients undergoing surgery.

Conditional survival (CS) is a statistical measure that estimates the likelihood of survival for a specific duration after diagnosis or treatment, considering the patient’s current survival time^11^. This approach provides a more personalized prognosis that evolves over time, making it more relevant and meaningful to patients than the traditional survival analysis. It allows physicians to tailor checkup schedules and content based on a patient’s unique survival pattern, enabling personalized adjustments. Previous studies on conditional survival in colorectal cancer have been conducted^12–14^, but most have included a diverse age range, and few have focused exclusively on the older population. A nomogram, which is a predictive tool for prognosis, has emerged as valuable in assessing the chances of survival in elderly colon cancer patients^15^. However, the nomogram overlooks the crucial aspect of incorporating patient survival duration. This study aimed to investigate conditional survival rates in elderly non-metastatic colon cancer patients who underwent colectomy, while developing nomograms to assess their prognosis.

## 2. Materials and methods

### Data collection

This study used data from the November 2020 release of the Surveillance, Epidemiology, and End Results (SEER) database obtained specifically from SEER*Stat Version 8.4.2. The study adhered to the guidelines of the 1964 Declaration of Helsinki and was conducted according to the STROCSS criteria. As per our institute’s Ethics Committee, this study did not require approval from an institutional review board because it used de-identified public database information. The study included a subset of elderly colon cancer patients who underwent colectomy and met the following criteria: (1) aged 70–89 years, (2) pathologically diagnosed with stage I-III colon cancer, (3) had undergone colectomy, and (4) had colon cancer as the only primary cancer. Patients who received neoadjuvant therapy were excluded from the analysis because of their potential effects on downstaging and prognostic factors. Patients without clinicopathological data or survival information were excluded from this study. A diagram outlining the screening process is presented in Supplementary Figure 1.

### Statistical analyses

In this study, the time from diagnosis to death from any cause was referred to as overall survival (OS), while the time from diagnosis to death, specifically from colon cancer, was referred to as colon-specific survival (CSS). A multivariate Cox regression model was constructed to assess the correlation between clinicopathological characteristics and survival outcomes. Survival curves were generated using the Kaplan-Meier method, and survival disparities were assessed using the log-rank test. CS represents the likelihood of survival for an additional specified time period (y years) after survival for a certain duration (x years). This measure was derived from Kaplan-Meier survival data and was mathematically expressed as CS(x|y) = S(x+y)/S(x), where S(x) represents the OS or CSS rate at x years, as estimated the Kaplan-Meier method^16^. This study employed COS (x) and CCSS (x) as alternatives to S(x) to determine the number of survivors in year x. The standardized differences (d) method was used to compute variations in CS across the subgroups^17^. When |d| was less than 0.1, it indicated negligible difference within each group; when it ranged from 0.1 to less than 0.3, it signified a minor difference; when it falled between 0.3 and less than 0.5, it suggested a moderate difference; and when |d| was 0.5 or greater, it indicated a notable difference.

The study participants were randomly allocated to either the training or validation cohort with a distribution ratio of 7:3. Univariate Cox regression analysis was performed to identify potential variables for inclusion in multivariate analysis, with a requirement for significance of p<0.05. Subsequently, variables demonstrating prognostic significance in multivariate Cox regression analysis were used to construct nomograms. To evaluate the discriminative ability of the nomogram, the area under the receiver operating characteristic (ROC) curve (AUC) was calculated. The calibration assessment was conducted using the Brier score and a calibration plot, employing the bootstrapping approach with 1000 resampling bootstraps. After the nomogram was subjected to decision curve analysis (DCA), its clinical efficacy was assessed.

R version 4.1.1 was employed for statistical analysis. Categorical variables are presented as frequencies and percentages, and disparities among various groups were examined using the chi-squared test. Continuous variables were expressed as means with standard deviations and compared using the t-test or rank-sum test based on data distribution. Statistical significance was determined using a p-value less than 0.05.

## 3. Results

### Clinicopathological characteristics

This study encompassed 9302 cases from 2004 to 2017 based on specific criteria. As shown in Table 1, the training set consisted of 6511 patients, whereas the validation set comprised 2791 patients. The patients had an average age of 78.6 years with a standard deviation of 5.42. The majority of these patients were female (57.4%), white (81.0%), diagnosed after 2010 (55.2%), and had tumors located in the proximal colon (73.1%). Grade II patients constituted the largest proportion (70.2%), followed by grade III (19.7%), grade I (6.64%), and grade IV (3.48%) patients. Colon adenocarcinoma was the most common cancer type (89.7%). Tumors < 5 cm accounted for 58.0% of the cases, and the majority of patients were married (51.6%) and had negative or normal carcino-embryonic antigen (CEA) test results (62.6%). Tumor stages were classified as T1 (7.99%), T2 (16.7%), T3 (61.2%), and T4 (14.1%). Nodal stages were classified as N0 (63.4%), N1 (24.6%), and N2 (12.0%). A small proportion of the patients received radiotherapy (0.8%) or chemotherapy (22.5%). The training and validation cohorts exhibited no significant differences in any of the variables.

**Table 1.** Baseline characteristics of enrolled patients.

|  | Total ( N=9302) | Train (N=6511) | Test (N=2791) | p |
| --- | --- | --- | --- | --- |
| Age | 78.6 (5.42) | 78.6 (5.44) | 78.6 (5.40) | 0.894 |
| Race |  |  |  | 0.360 |
| White | 7533 (81.0%) | 5293 (81.3%) | 2240 (80.3%) |  |
| Black | 532 (5.72%) | 359 (5.51%) | 173 (6.20%) |  |
| Others | 1237 (13.3%) | 859 (13.2%) | 378 (13.5%) |  |
| Sex: |  |  |  | 0.744 |
| Male | 3964 (42.6%) | 2767 (42.5%) | 1197 (42.9%) |  |
| Female | 5338 (57.4%) | 3744 (57.5%) | 1594 (57.1%) |  |
| Year of Diagnosis: |  |  |  | 0.643 |
| < 2010 | 4165 (44.8%) | 2926 (44.9%) | 1239 (44.4%) |  |
| ≥ 2010 | 5137 (55.2%) | 3585 (55.1%) | 1552 (55.6%) |  |
| Site: |  |  |  | 0.100 |
| Proximal | 6802 (73.1%) | 4803 (73.8%) | 1999 (71.6%) |  |
| Distal | 2383 (25.6%) | 1629 (25.0%) | 754 (27.0%) |  |
| Others | 117 (1.26%) | 79 (1.21%) | 38 (1.36%) |  |
| Grade: |  |  |  | 0.689 |
| Grade I | 618 (6.64%) | 436 (6.70%) | 182 (6.52%) |  |
| Grade II | 6532 (70.2%) | 4549 (69.9%) | 1983 (71.0%) |  |
| Grade III | 1828 (19.7%) | 1299 (20.0%) | 529 (19.0%) |  |
| Grade IV | 324 (3.48%) | 227 (3.49%) | 97 (3.48%) |  |
| Histology Type: |  |  |  | 0.199 |
| Adenocarcinoma | 8343 (89.7%) | 5834 (89.6%) | 2509 (89.9%) |  |
| Mucinous adenocarcinoma | 769 (8.27%) | 533 (8.19%) | 236 (8.46%) |  |
| Others | 190 (2.04%) | 144 (2.21%) | 46 (1.65%) |  |
| Tumor Size: |  |  |  | 0.961 |
| < 5 cm | 5394 (58.0%) | 3774 (58.0%) | 1620 (58.0%) |  |
| ≥ 5 cm | 3908 (42.0%) | 2737 (42.0%) | 1171 (42.0%) |  |
| Marital Status: |  |  |  | 0.305 |
| Married | 4803 (51.6%) | 3372 (51.8%) | 1431 (51.3%) |  |
| Single | 831 (8.93%) | 566 (8.69%) | 265 (9.49%) |  |
| Widowed | 2892 (31.1%) | 2045 (31.4%) | 847 (30.3%) |  |
| Separated/Divorced | 776 (8.34%) | 528 (8.11%) | 248 (8.89%) |  |
| CEA: |  |  |  | 0.535 |
| Negative/Normal | 5825 (62.6%) | 4091 (62.8%) | 1734 (62.1%) |  |
| Positive/elevated | 3477 (37.4%) | 2420 (37.2%) | 1057 (37.9%) |  |
| T stage: |  |  |  | 0.282 |
| T1 | 743 (7.99%) | 513 (7.88%) | 230 (8.24%) |  |
| T2 | 1553 (16.7%) | 1086 (16.7%) | 467 (16.7%) |  |
| T3 | 5697 (61.2%) | 4022 (61.8%) | 1675 (60.0%) |  |
| T4 | 1309 (14.1%) | 890 (13.7%) | 419 (15.0%) |  |
| N stage: |  |  |  | 0.576 |
| N0 | 5899 (63.4%) | 4141 (63.6%) | 1758 (63.0%) |  |
| N1 | 2290 (24.6%) | 1606 (24.7%) | 684 (24.5%) |  |
| N2 | 1113 (12.0%) | 764 (11.7%) | 349 (12.5%) |  |
| Radiation |  |  |  | 0.177 |
| None/Unknown | 9228 (99.2%) | 6465 (99.3%) | 2763 (99.0%) |  |
| Yes | 74 (0.80%) | 46 (0.71%) | 28 (1.00%) |  |
| Chemotherapy: |  |  |  | 0.468 |
| None/Unknown | 7206 (77.5%) | 5030 (77.3%) | 2176 (78.0%) |  |
| Yes | 2096 (22.5%) | 1481 (22.7%) | 615 (22.0%) |  |

### COS and CCSS

Figure 1 shows the conditional survival curves. The analysis demonstrated a consistent pattern of improvement in the actuarial OS and CSS of individuals as the duration of survival increased annually. Meanwhile, as shown in Table 2 and Figure 1, the COS5 rates initially saw a marginal uptick, followed by a subsequent decline. Specifically, the rates at baseline, one year, two years, three years, four years, and five years post-diagnosis were 62.5%, 63.2%, 62.8%, 62.1%, 61.6%, and 59.8%, respectively. However, the CCSS5 rates exhibited an upward trajectory over the five-year period, with the initial and subsequent conditional survival rates at baseline and years 1 through 5 being 78.1%, 80.9%, 84.2%, 86.9%, 89.3%, and 90.9%, respectively (Table 3).

**Figure 1.**
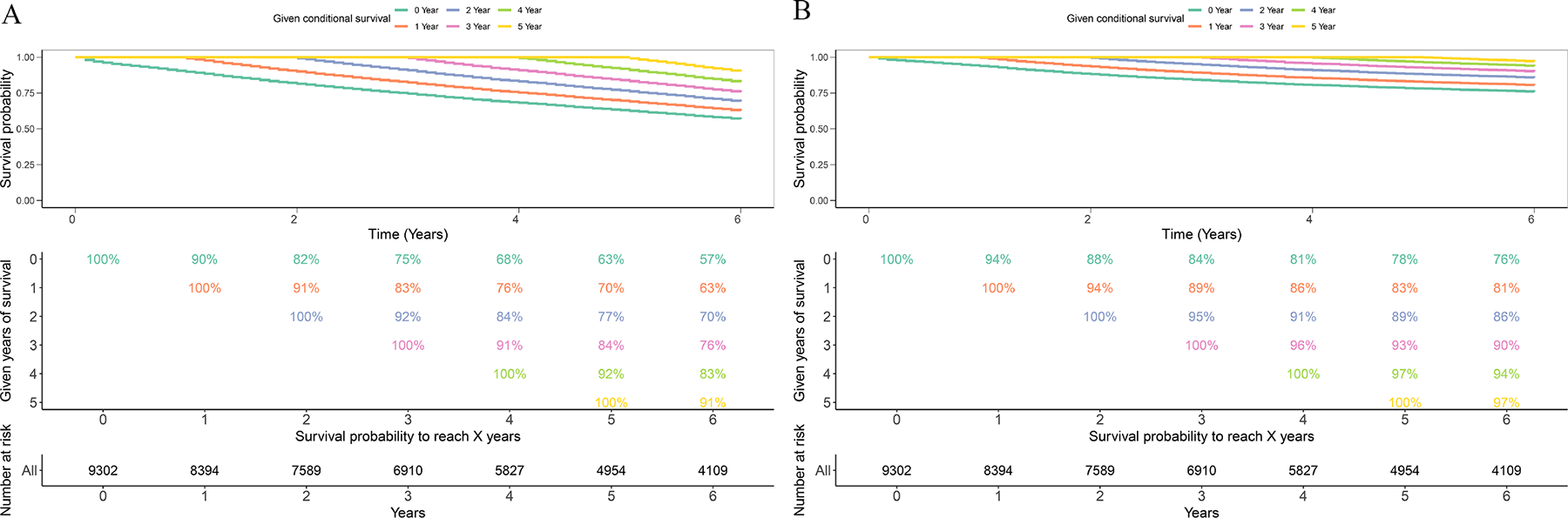
Conditional survival analysis of elderly non-metastatic colon cancer patients who undergo colectomy. (A) Kaplan–Meier curves estimating overall survival for individuals who had successfully survived a duration of 0-6 years. (B) Kaplan–Meier curves estimating colon-specific survival for individuals who had successfully survived a duration of 0-6 years.

**Table 2.**
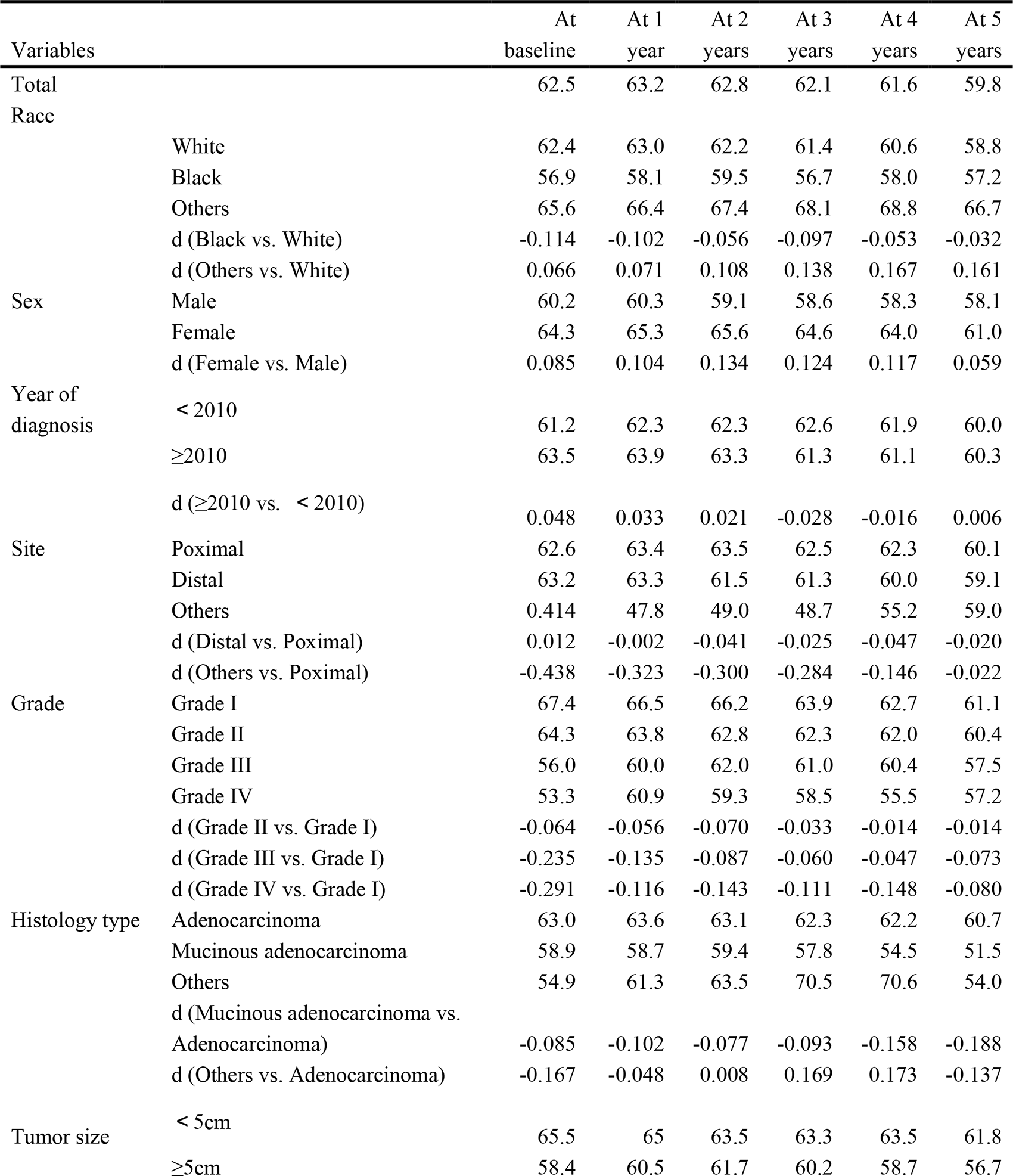
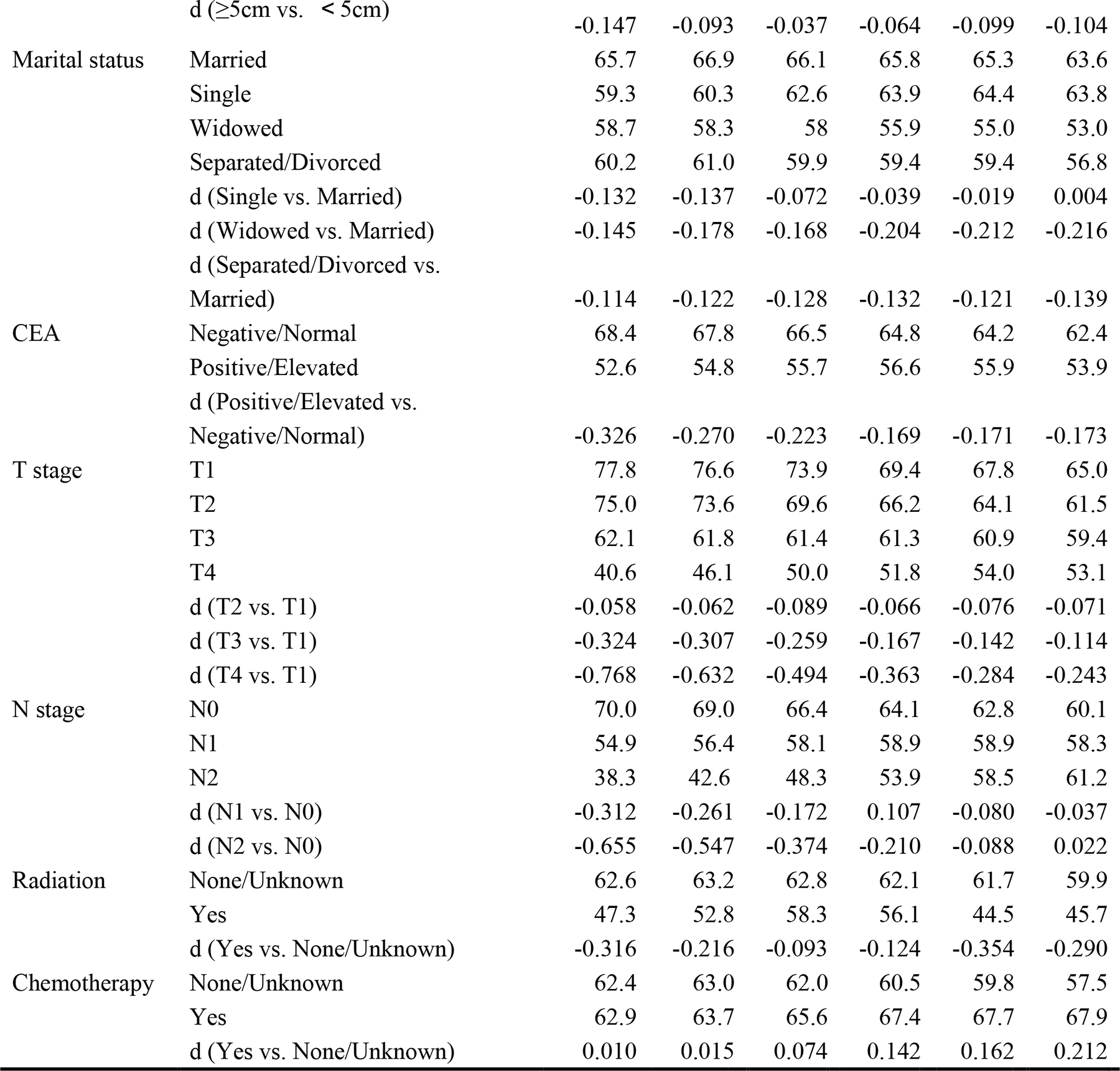
Five-year conditional overall survival rates (%).

**Table 3.**
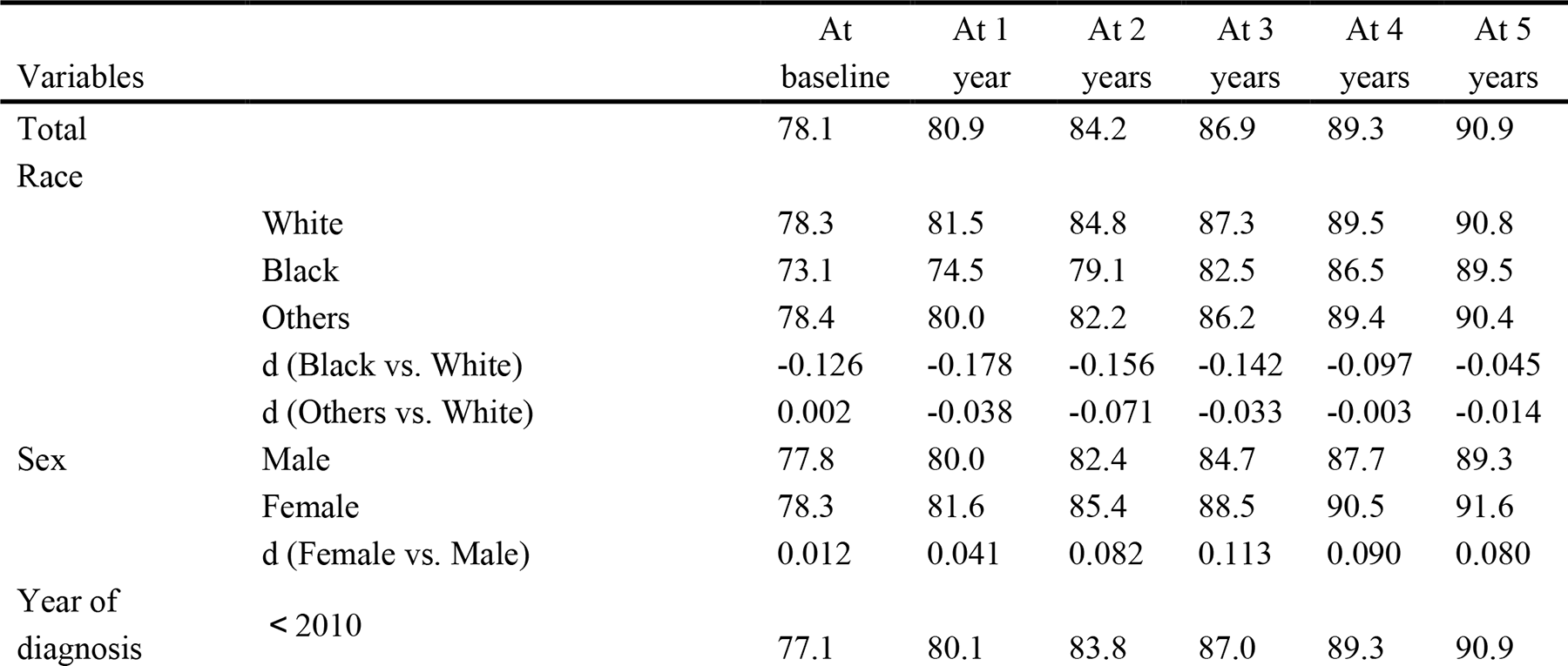
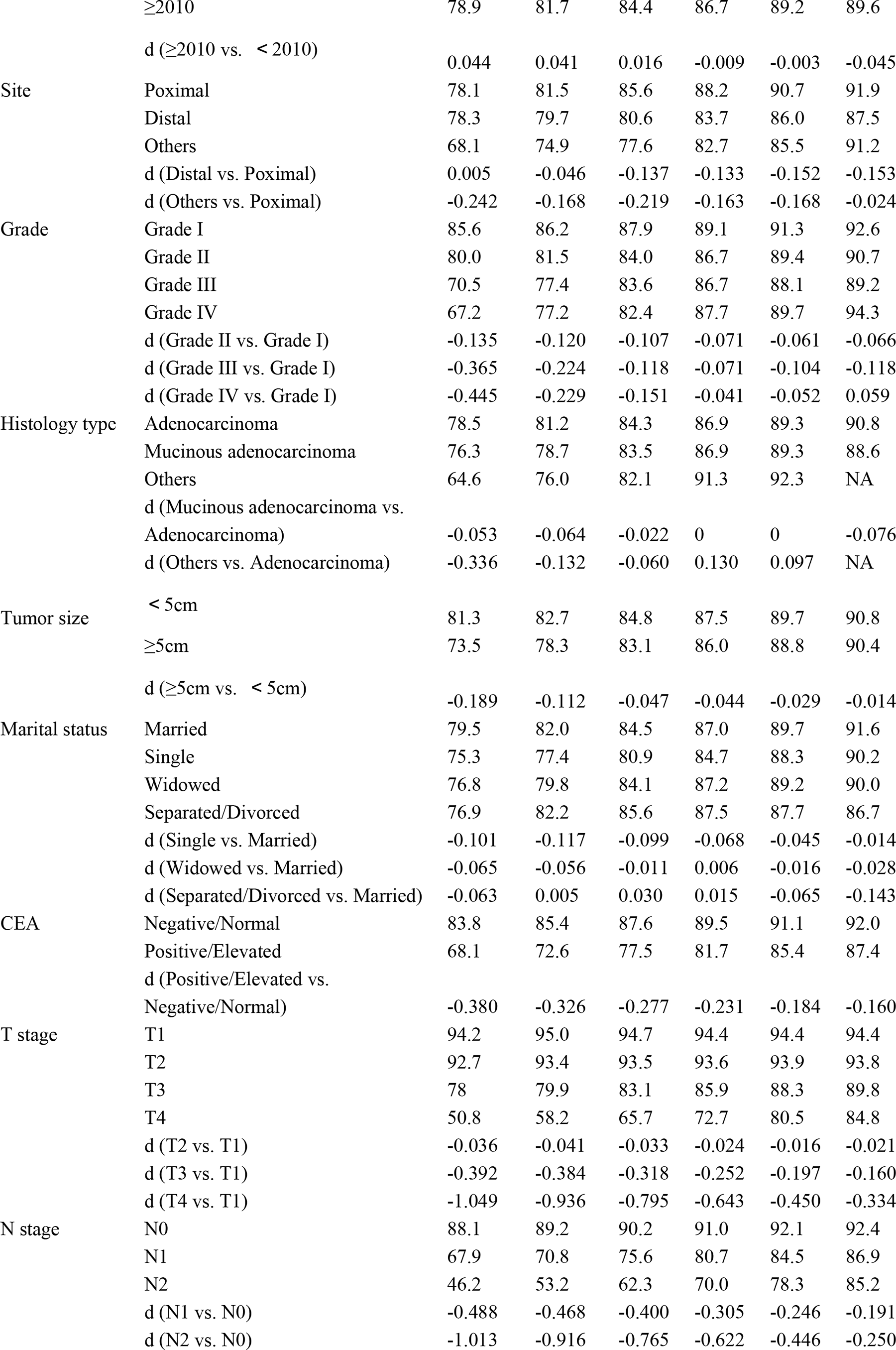
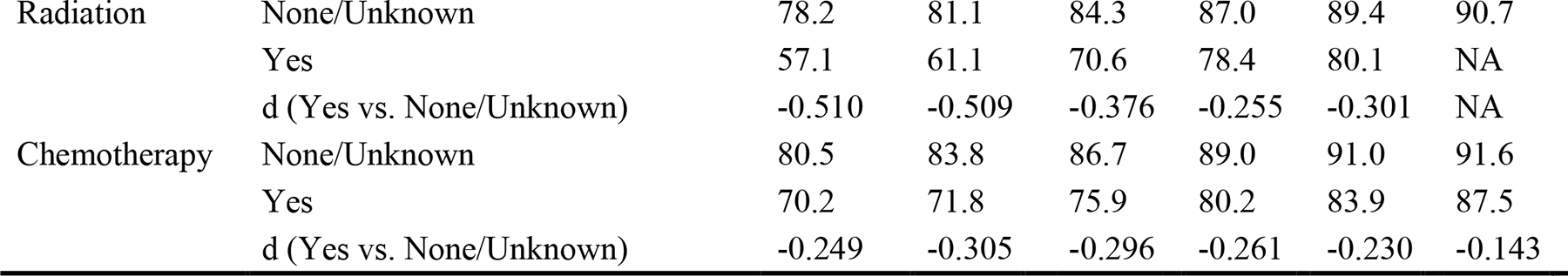
Five-year conditional colon-specific survival rates (%).

Subsequently, detailed subgroup analysis was conducted to assess the impact of various factors on the rates of COS5 and CCSS5. The expression rates of COS5 and CCSS5 varied across these subgroups, displaying distinct patterns over time. For COS5, there were persistent negligible or minor differences in survival between some subgroups (|d|<0.1: ≥2010 vs.<2010, Distal vs. Poximal, Grade II vs. Grade I, and T2 vs. T1; 0.1<|d|<0.3: Widowed vs. Married and Separated/Divorced vs. Married). The differences in COS5 levels between some subgroups gradually appeared over time (|d|<0.1→ |d|>0.1: Others vs. White and Chemotherapy Yes vs. None/Unknown). The disparity in COS5 rates among some subgroups slowly diminished over time (|d|>0.1→ |d|<0.1, |d|>0.3→ |d|<0.3, or |d|>0.5→ |d|<0.5; black vs. White, Others vs. proximal, grade III vs. Grade I, Grade IV vs. Grade I, Single vs. Married, Positive/Elevated vs. Negative/Normal, T3 vs. T1, T4 vs. T1, N1 vs. N0, and N2 vs. N0). Over time, the variance in the COS5 rate fluctuated irregularly across other subgroups. Correspondingly, for CCSS5, there were persistent negligible differences in survival between some subgroups (|d|<0.1: others vs. white, ≥ 2010 vs. < 2010, mucinous adenocarcinoma vs. adenocarcinoma, owed vs. Married and T2 vs. T1). The variance in CCSS5 among the subgroups became increasingly evident as time progressed (|d|<0.1→ |d|>0.1: Distal vs. Poximal). Over time, there was a gradual reduction in the differences in CCSS5 rates within certain subgroups (|d|>0.1→ |d|<0.1, |d|>0.3→ |d|<0.3, or |d|>0.5→ |d|<0.5: black vs. White, Grade II vs. Grade I, Grade IV vs. Grade I, ≥5 cm vs. < 5 cm, single vs. Married, Positive/Elevated vs. Negative/Normal, T3 vs. T1, T4 vs. T1, N1 vs. N0, and N2 vs. N0). The differences in the CCSS5 rate among the other subgroups fluctuated irregularly over time.

### Variables linked to the OS and CSS in patients who have attained a defined duration of survival

Age, race, sex, year of diagnosis, site, grade, marital status, CEA level, T stage, N stage, and chemotherapy were independent prognostic factors for OS at baseline (Table 4 and Supplementary Table 1). The prognostic factors for patients who survived for at least one year differed from those at baseline, including age, race, sex, marital status, CEA levels, T stage, N stage, and chemotherapy. After three years, age, race, sex, histology type, marital status, CEA levels, T stage, N stage, and chemotherapy independently affected OS. At the 5-year mark, age, race, sex, marital status, CEA levels, T stage, and chemotherapy were key predictors of OS, consistent with the baseline and other survival intervals.

**Table 4.** Multivariate analyses of overall deaths in baseline, ≥ one-, three-, five-year survivors in the training cohort.

| Variables |  | Baseline survivors |  | > 1 year survivors |  | > 3 year survivors |  | > 5 year survivors |  |
| --- | --- | --- | --- | --- | --- | --- | --- | --- | --- |
|  |  | HR | p | HR | p | HR | p | HR | p |
| Age |  | 1.07 (1.06-1.08) | <0.001 | 1.08 (1.07-1.08) | <0.001 | 1.09 (1.08-1.10) | <0.001 | 1.10 (1.09-1.12) | <0.001 |
| Race | White | Ref |  | Ref |  | Ref |  |  |  |
|  | Black | 1.20 (1.04-1.38) | 0.010 | 1.10 (0.95-1.28) | 0.217 | 1.06 (0.87-1.29) | 0.579 | 1.14 (0.90-1.44) | 0.287 |
|  | Others | 1.20 (1.04-1.38) | <0.001 | 0.77 (0.69-0.85) | <0.001 | 0.77 (0.67-0.87) | <0.001 | 0.69 (0.62-0.77) | <0.001 |
| Sex | Male | Ref |  | Ref |  | Ref |  |  |  |
|  | Female | 0.73 (0.68-0.78) | <0.001 | 0.71 (0.66-0.77) | <0.001 | 0.67 (0.61-0.74) | <0.001 | 0.69 (0.62-0.77) | <0.001 |
| Year of diagnosis | < 2010 |  |  |  |  |  |  |  |  |
| | ≥2010 | 0.90 (0.84-0.98) | 0.004 | | | | | | |
| Site | Poximal | Ref |  |  |  |  |  |  |  |
|  | Distal | 1.04 (0.97-1.12) | 0.346 |  |  |  |  |  |  |
|  | Others | 1.52 (1.16-2.00) | 0.003 |  |  |  |  |  |  |
| Grade | Grade I | Ref |  | Ref |  |  |  |  |  |
|  | Grade II | 1.04 (0.91-1.19) | 0.537 | 0.96 (0.83-1.10) | 0.524 |  |  |  |  |
|  | Grade III | 1.17 (1.01-1.35) | 0.035 | 0.97 (0.83-1.13) | 0.672 |  |  |  |  |
|  | Grade IV | 1.21 (0.98-1.50) | 0.074 | 0.97 (0.76-1.24) | 0.672 |  |  |  |  |
| Histology type | Adenocarcinoma | Ref |  | Ref |  | Ref |  | Ref |  |
|  | Mucinous adenocarcinoma | 1.04 (0.93-1.16) | 0.462 | 1.04 (0.92-1.18) | 0.494 | 1.06 (0.92-1.23) | 0.393 | 1.09 (0.92-1.30) | 0.311 |
|  | Others | 0.81 (0.64-1.01) | 0.064 | 0.90 (0.68-1.18) | 0.452 | 0.65 (0.45-0.95) | 0.025 | 0.80 (0.52-1.22) | 0.297 |
| Tumor size | < 5cm | Ref |  | Ref |  |  |  | Ref |  |
| | ≥5cm | 1.01 (0.94-1.08) | 0.794 | 0.96 (0.89-1.04) | 0.324 | | | 1.03 (0.92-1.14) | 0.608 |
| Marital status | Married | Ref |  | Ref |  | Ref |  | Ref |  |
|  | Single | 1.09 (0.97-1.23) | 0.152 | 1.12 (0.98-1.28) | 0.089 | 1.21 (1.03-1.41) | 0.02 | 1.02 (0.83-1.24) | 0.874 |
|  | Widowed | 1.14 (1.05-1.23) | 0.001 | 1.15 (1.05-1.25) | 0.002 | 1.24 (1.12-1.37) | <0.001 | 1.22 (1.08-1.38) | 0.002 |
|  | Separated/Divorced | 1.45 (1.29-1.64) | <0.001 | 1.36 (1.19-1.56) | <0.001 | 1.47 (1.25-1.73) | <0.001 | 1.57 (1.29-1.92) | <0.001 |
| CEA | Negative/Normal | Ref |  |  |  | Ref |  |  |  |
|  | Positive/Elevated | 1.38 (1.29-1.47) | <0.001 | 1.35 (1.25-1.45) | <0.001 | 1.39 (1.26-1.50) | <0.001 | 1.26 (1.13-1.40) | <0.001 |
| T stage | T1 | Ref |  | Ref |  | Ref |  | Ref |  |
|  | T2 | 1.03 (0.88-1.20) | 0.746 | 0.99 (0.84-1.16) | 0.880 | 1.08 (0.90-1.29 ) | 0.419 | 1.06 (0.86-1.31) | 0.586 |
|  | T3 | 1.21 (1.05-1.39) | 0.009 | 1.19 (1.03-1.38) | 0.022 | 1.12 (0.95-1.32) | 0.161 | 1.07 (0.88-1.30) | 0.519 |
|  | T4 | 2.02 (1.72-2.38) | <0.001 | 1.84 (1.54-2.18) | <0.001 | 1.42 (1.16-1.73) | <0.001 | 1.31 (1.02-1.70) | 0.037 |
| N stage | N0 | Ref |  | Ref |  | Ref |  |  |  |
|  | N1 | 1.47 (1.35-1.59) | <0.001 | 1.51 (1.38-1.66) | <0.001 | 1.26 (1.13-1.41) | <0.001 |  |  |
|  | N2 | 2.30 (2.06-2.56) | <0.001 | 2.27 (2.00-2.57) | <0.001 | 1.64 (1.40-1.93) | <0.001 |  |  |
| Chemotherapy | None/Unknown | Ref |  | Ref |  | Ref |  | Ref |  |
|  | Yes | 0.65 (0.59-0.71) | <0.001 | 0.65 (0.59-0.73) | <0.001 | 0.79 (0.70-0.90) | <0.001 | 0.83 (0.72-0.96) | 0.009 |

HR Hazard ratio

Age, year of diagnosis, site, grade, marital status, CEA, T stage, N stage, and chemotherapy were determined to be independent prognostic factors for cancer-specific survival (CSS) at baseline, as indicated in Table 5 and Supplementary Table 2. After a minimum of one year of survival, age, CEA levels, T stage, N stage, and chemotherapy remained significant risk factors for CSS. For patients who survived beyond three years, age, sex, tumor location, marital status, CEA levels, T stage, and N stage emerged as influential factors. Among those who surpassed a five-year milestone, age, tumor site, CEA level, and T stage were independent prognostic factors. Constant predictors of CSS for newly diagnosed patients and 1-, 3-, and 5-year survivors were age, CEA, and T stage.

**Table 5.**
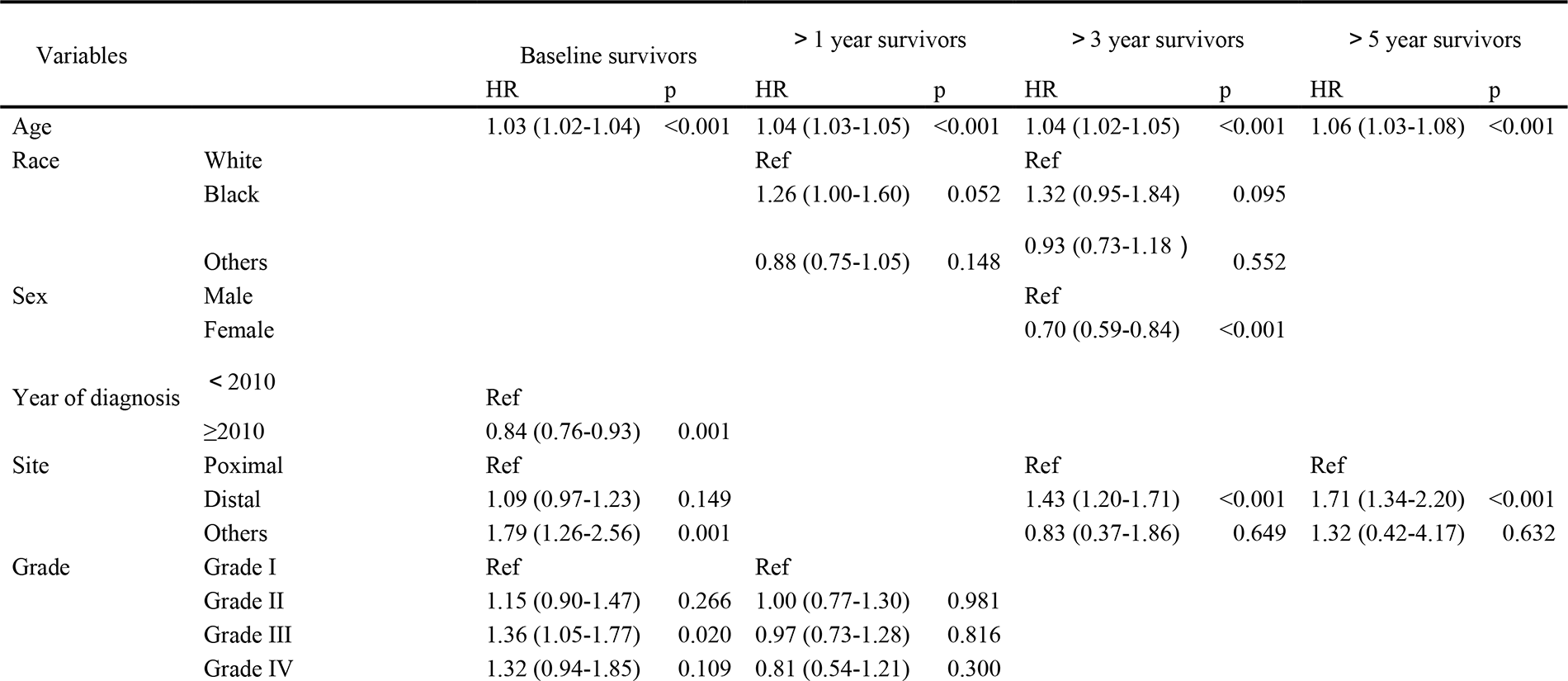
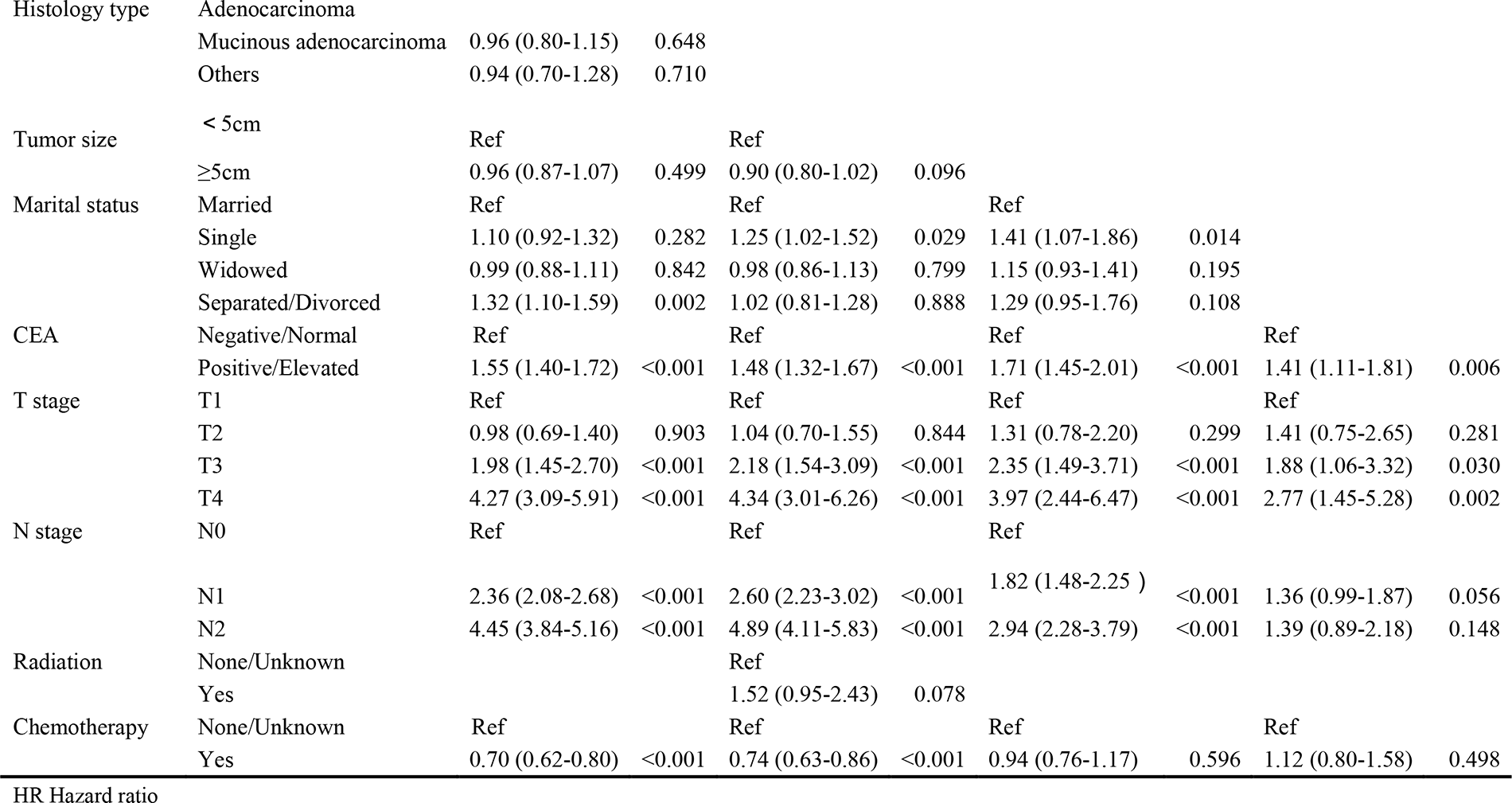
Multivariate analyses of colon-specific deaths in baseline, ≥ one-, three-, five-year survivors in the training cohort.

### Conditional nomogram construction and validation

Figure 2 and 3 illustrate the OS and CSS nomograms developed for cancer patients at different stages of their journey, including the start and at the one-year, three-year, and five-year milestones. These nomograms consider the specific risk factors for each individual. Our investigation delved deeper into the performances of these nomograms. Baseline analysis of the OS nomogram in the training set revealed an area under the curve (AUC) of 0.703, 0.713, and 0.713 for the one-year, three-year, and five-year intervals, respectively (Figure 4). In the test set, the corresponding AUC were 0.714, 0.722, and 0.719, respectively (Supplementary Figure 2). For patients who survived for at least one year, the OS nomogram showed AUC of 0.700, 0.697, and 0.698 in the training set and 0.698, 0.694, and 0.701 in the test set, respectively, for the same intervals. For patients surviving for 3 years, the OS nomogram achieved AUC of 0.652, 0.665, and 0.694 in the training set and 0.663, 0.666, and 0.688 in the test set, respectively. Additionally, for patients surviving 5 years, the training set’s OS nomogram yielded AUC of 0.661, 0.694, and 0.695 for the one-year, three-year, and five-year durations. In the test set, AUC were 0.590, 0.674, and 0.681 for the same intervals. Regarding the CSS nomogram, at baseline, the training cohort demonstrated AUC values of 0.771, 0.788, and 0.782 for the one-year, three-year, as well as five-year periods (Figure 5), while the test cohort achieved corresponding AUC values of 0.787, 0.804, and 0.800, respectively (Supplementary Figure 3). For patients who survived for 1 year, the training cohort CSS nomogram attained AUC of 0.785, 0.795, and 0.781 for the one-year, three-year, and five-year intervals, whereas the test cohort displayed AUC of 0.780, 0.778, and 0.761, respectively. Among patients surviving 3 years, the training cohort showed AUC of 0.765, 0.732, and 0.727 over the one-year, three-year, and five-year periods, whereas the test cohort displayed AUC of 0.786, 0.732, and 0.697, respectively, for the same duration. For patients who survived for 5 years, the training cohort CSS nomogram achieved AUC values of 0.664, 0.669, and 0.682 for the one-year, three-year, and five-year periods, whereas the test cohort showed AUC values of 0.585, 0.629, and 0.655, respectively.

**Figure 2.**
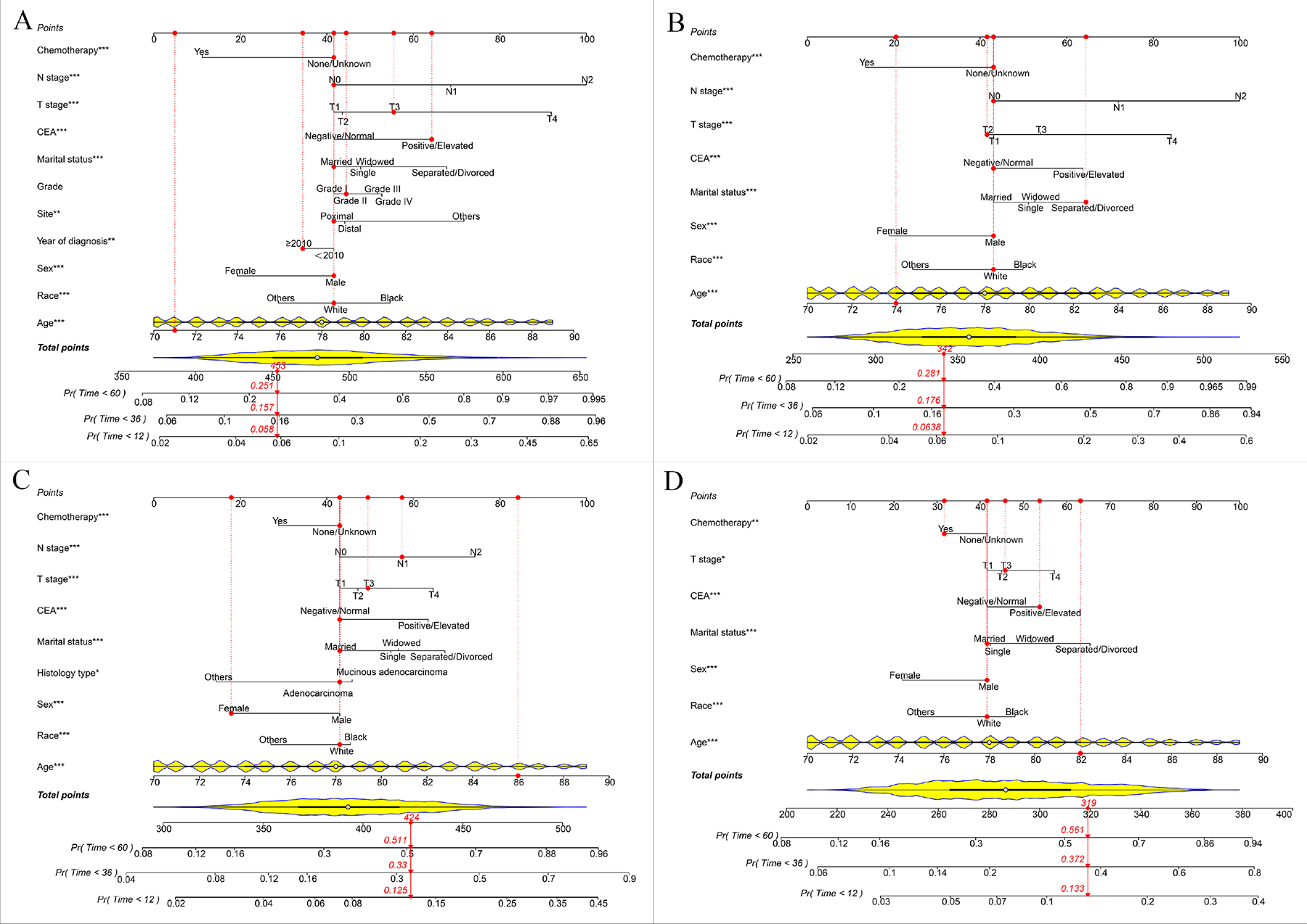
Nomograms to predict one-, three-, and five-overall survival for baseline (A), one-year (B), three-year (C), and five-year (D) survivors.

**Figure 3.**
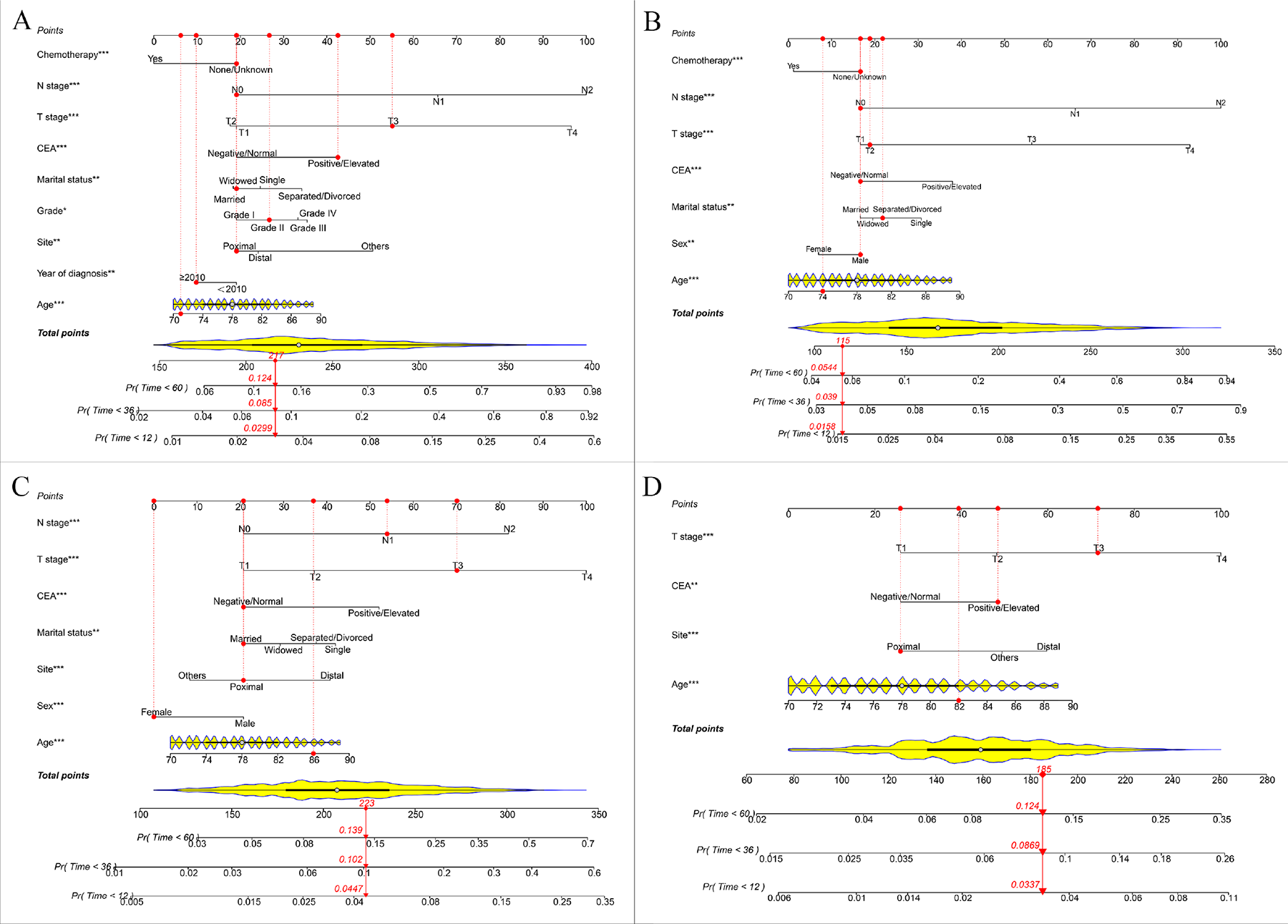
Nomograms to predict one-, three-, and five-year colon-specific survival for baseline (A), one-year (B), three-year (C), and five-year (D) survivors.

**Figure 4.**
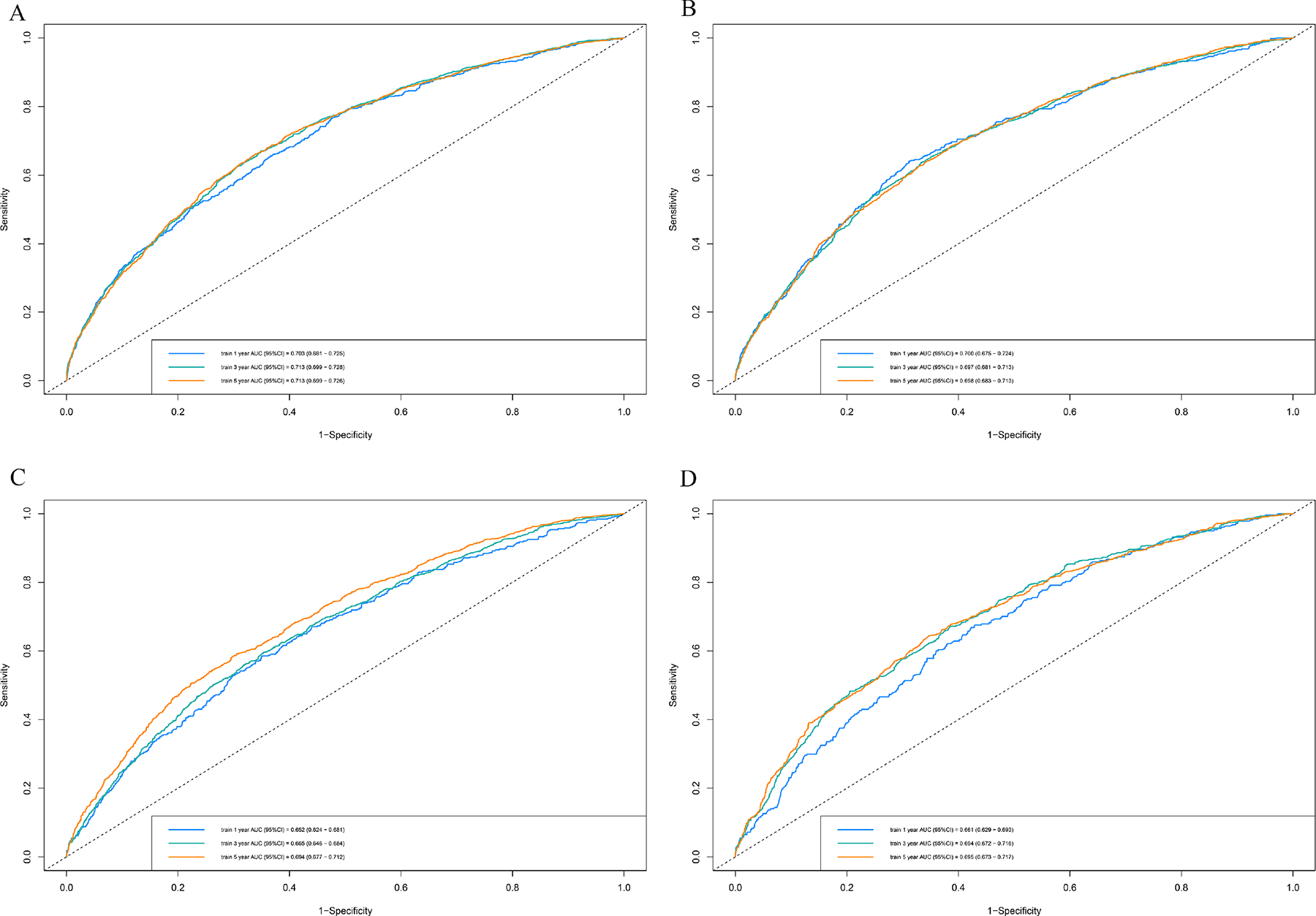
ROC curve analyses in the training cohort for one-, three-, and five-year overall survival in baseline (A), one-year (B), three-year (C), and five-year (D) survivors. ROC, receiver operator characteristic curve; AUC, area under the curve.

**Figure 5.**
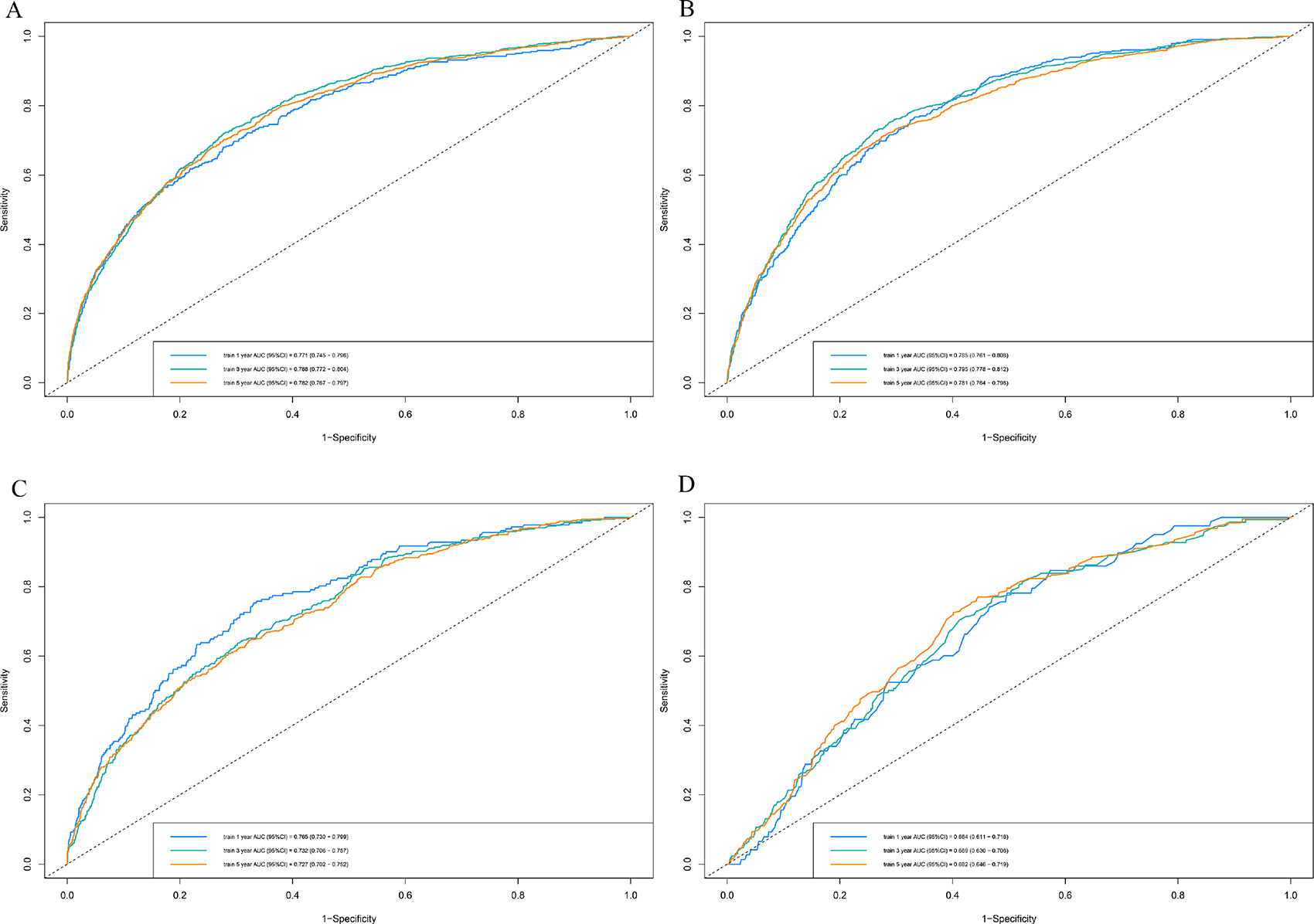
ROC curve analyses in the training cohort for one-, three-, and five-year colon-specific survival in baseline (A), one-year (B), three-year (C), and five-year (D) survivors. ROC, receiver operator characteristic curve; AUC, area under the curve.

The calibration curves of the OS and CCS nomograms when utilized for these training and validation sets are also presented. The outcomes demonstrated strong concordance between anticipated and observed survival rates in both the training and validation sets, indicating the accuracy of these nomograms in predicting survival probabilities (Supplementary Figure 4-7). In addition, the DCA curves also indicated the favorable clinical utility of these nomograms (Supplementary Figure 8-11).

## 4. Discussion

In this investigation, we examined the survival data of elderly patients with non-metastatic colon cancer who had undergone colectomy in detail. The uniqueness and notable attributes of our research encompass the following: we conducted an analysis of the CS rates, allowing for a more precise assessment of OS and CSS outcomes in both medium-term and long-term survivors; this analysis aids in accurately forecasting survival rates and advancing personalized medical approaches; our findings revealed the nomograms to possess favorable discriminatory and predictive capabilities when forecasting OS and CSS rates.

In the coming years, we expect a higher incidence of colon cancer in the elderly population. This can be attributed to a longer life expectancy and improved rates of survival^18^. Colectomy is the primary curative treatment for the elderly. Previous studies have reported a notable correlation between surgical resection and improved prognosis in older patients with colon cancer^8–9,19^. However, the prognosis of elderly patients differs from that of younger individuals owing to the presence of other health conditions, functional disabilities, more advanced tumors, and a decreased ability to maintain internal balance. Elderly patients experience higher rates of complications and mortality following surgery as well as a less favorable overall outlook^3,20^. Given the complexities surrounding older adults with colon cancer, there is an urgent need for further research focusing on this demographic, particularly on those who have undergone surgical procedures. Currently, there is limited information on the medium- and long-term postoperative prognoses of elderly patients with colon cancer, and few studies have investigated the practical application of prediction models specifically tailored to this population. Wang^21^ et al. developed a prognostic model that accurately predicted the outcomes of elderly patients with colorectal cancer. However, their study included patients with and without metastatic disease, and it is widely recognized that prognosis differs significantly between these two groups. Additionally, their study focused only on CSS and not OS. To our knowledge, this study is the first to investigate the development and validation of a reliable nomogram for predicting both OS and CSS in elderly adults with non-metastatic colon cancer who have undergone colectomy. This model is expected to provide patients and healthcare providers with a valuable tool for guiding follow-up care and treatment decisions.

CS provides a precise estimation of the likelihood of a specific time point in the course of a disease, closely tied to individual long-term survival rates based on patients’ time of survival^11^. This allows for a more accurate depiction of prognostic outcomes. To our knowledge, this study is the first to show CS in elderly individuals with non-metastatic colon cancer who have undergone colectomy. In this study, survival analysis revealed an upward trend in actuarial OS and CSS among elderly patients as their survival duration increased annually. However, COS and CCSS rates showed contrasting patterns. Initially, the COS5 rates experienced a slight increase, followed by a subsequent decrease, whereas the CCSS5 rates demonstrated a consistent upward trend over the five-year duration. Compared with the previously noted trend of annual increases in the likelihood of COS rates for malignant tumors^11,13^, particularly CRC, this study revealed a distinctive pattern of change in COS rates. Our hypothesis attributed this variance to disparity in the cohort composition. This study differs from previous research by exclusively focusing on older adults instead of encompassing individuals across all age groups. Throughout the course of follow-up, the impact of non-colon cancer-related death determinants accumulated at various stages of the life course, leading to a growing risk of other conditions such as cardiovascular and cerebrovascular diseases among elderly patients with colon cancer. a recent study^22^, it was found that the majority of deaths in patients aged > 80 years during the follow-up period were unrelated to CRC. Non-CRC-related causes of death, such as respiratory failure, particularly aspiration pneumonia, cardiovascular disease, and sepsis, were the primary contributors to mortality. Another study suggested that cerebrovascular-specific hazard ratios were 3.10-, 6.67, and 10.95 times greater for CRC patients aged 50-64, 65-74, and over 75 years, respectively, compared to those under 50 years old^23^. One of the foremost concerns for those who survive cancer is the risk of mortality. Although the risk of death due to colon cancer surpasses that of other illnesses in the initial years, this specific risk diminishes over time in a discernible trend. Our research showed that over the span of five years, the CCSS5 showed a consistent increase, with the baseline and subsequent conditional survival rates at years 1 through 5 documented as 78.1%, 80.9%, 84.2%, 86.9%, 89.3%, and 90.9%, respectively, indicating a positive trend. The findings from the CS analysis could instill greater optimism in the patients. For instance, consider a patient who initially has a 5-year CSS of 78.1% at the time of diagnosis. Subsequently, through dedicated adherence to treatment and follow-up, the patient successfully achieved 5-year survival, and the CS analysis conveyed the encouraging news: “Your 5-year CSS rate has risen to 90.9%.” On the other hand, in contrast to the general population’s COS outcomes, the comparatively less positive COS outcomes in the elderly indicate a greater need for physicians to prioritize non-colonic causes of death as patients age. Subgroup analysis was performed to further understand the influence of different variables on CS. The rates of COS5 and CCSS5 expression varied in these subgroups. Over time, the variation in the rates of both COS5 and CCSS5 among many subgroups gradually decreased (black vs. White, Grade IV vs. Grade I, Single vs. Married, Positive/Elevated vs. Negative/Normal, T3 vs. T1, T4 vs. T1, N1 vs. N0, and N2 vs. N0). One possible explanation is that patients in each subgroup with some high-risk factors died over time, whereas the remaining patients had a better prognosis, resulting in a smaller difference in survival between the subgroups. The change patterns of COS5 and CCSS5 differed among the subgroups, which may be influenced by multiple factors, such as those affecting cardiovascular death. Future studies of CS in these subgroups should be conducted.

Our study examined the impact of various baseline prognostic factors on patients with different survival durations. Multiple factors, including age, race, sex, year of diagnosis, site, grade, marital status, carcinoembryonic antigen (CEA) levels, T stage, N stage, and chemotherapy, significantly influenced OS according to multifactor analysis. Upon reaching the critical 5-year survival milestone, we identified several key independent prognostic factors that consistently influenced the patient outcomes. These factors included age, race, sex, marital status, CEA levels, T stage, and chemotherapy, demonstrating a predictive value for OS across baseline and 1-year, 3-year, and 5-year intervals. Furthermore, we observed that at the beginning of the study, age, year of diagnosis, site, grade, marital status, CEA levels, T stage, N stage, and chemotherapy independently influenced CSS. However, as time progressed, only age, CEA level, and T stage consistently emerged as predictive factors for CSS in both newly diagnosed patients and those who survived for 1, 3, and 5 years. These variables, as observed in previous studies, could predict OS and CSS at the beginning of diagnosis^15,19,21^, but their predictive significance diminished over time. To provide more accurate prognostic predictions, we utilized these variables to create nomograms for each type of survival, which demonstrated good prediction performance and a correspondingly good match between the predicted and observed survival rates in the calibration plots. The practical application of these nomograms is also supported by their favorable clinical utility, as shown in the DCA curves. However, it should be noted that, as time progressed, the AUC values of the nomograms gradually declined, indicating a decrease in predictive accuracy. This trend can be explained by the diminishing impact of clinicopathological factors on the prognosis over time. Further research is needed to assess the latest clinical and laboratory factors at specific time points to more accurately identify prognostic factors affecting elderly colon cancer survivors. This study had some limitations. First, given the retrospective nature of the study, it inevitably introduced a degree of selection bias owing to its inherent design. Second, obtaining postoperative complications and adjuvant treatment details from the SEER database for all patients is not feasible, potentially impacting the survival prognosis of elderly individuals with colon cancer. Third, the analysis of CS necessitates extensive data, thus posing challenges for the external validation of these nomograms. Furthermore, the ongoing update of nomograms is essential to accommodate the evolving treatment strategies.

## 5. Conclusion

We reported the rates of CS in elderly non-metastatic colon cancer patients following colectomy, enhancing our understanding of survival rates in the medium- and long-term. We also created personalized CS nomograms to predict mortality risk at different intervals. These tools aid surgeons and patients in making informed decisions, and contribute to personalized medicine.

## Data Availability

All data produced in the present study are available upon reasonable request to the authors.

## Ethical approval

The information used in our study was obtained from the SEER database, which is accessible to the public, thus eliminating the need for ethical clearance.

## Acknowledgments

The invaluable support of ChatGPT was instrumental in enhancing the sentence structure revisions.

---

**Supplementary Table 1.** Univariate analyses of overall deaths in baseline, ≥ one-, three- and five-year survivors in the training cohort.

**Supplementary Table 2.** Univariate analysis of colon-specific deaths at baseline, ≥ one-, three-and five-year survivors in the training cohort.

**Supplementary Figure 1.** Flow diagram for enrollment of the study cohort.

**Supplementary Figure 2.** ROC curve analyses in the test cohort for one-, three-, and five-year overall survival in baseline (A), one-year (B), three-year (C), and five-year (D) survivors. ROC, receiver operator characteristic curve; AUC, area under the curve.

**Supplementary Figure 3.** ROC curve analyses in the test cohort for one-, three-, and five-year colon-specific survival in baseline (A), one-year (B), three-year (C), and five-year (D) survivors. ROC, receiver operator characteristic curve; AUC, area under the curve.

**Supplementary Figure 4.** Calibration curves of baseline nomograms. (A, C and E) showed Calibration curves of nomograms in predicting one-, three-, and five-year OS, respectively. (B, D and F) showed Calibration curves of nomograms in predicting one-, three-, and five-year CSS, respectively. OS, overall survival; CSS, colon-specific survival.

**Supplementary Figure 5.** Calibration curves of one-year survivors nomograms. (A, C and E) showed Calibration curves of nomograms in predicting one-, three-, and five-year OS, respectively. (B, D and F) showed Calibration curves of nomograms in predicting one-, three-, and five-year CSS, respectively. OS, overall survival; CSS, colon-specific survival.

**Supplementary Figure 6.** Calibration curves of three-year survivors nomograms in the training cohort. (A, C and E) showed Calibration curves of nomograms in predicting one-, three-, and five-year OS, respectively. (B, D and F) showed Calibration curves of nomograms to predict one-, three-, and five-year CSS, respectively. OS, overall survival; CSS, colon-specific survival.

**Supplementary Figure 7.** Calibration curves of five-year survivors nomograms in the training cohort. (A, C and E) showed Calibration curves of nomograms in predicting one-, three-, and five-year OS, respectively. (B, D and F) showed Calibration curves of nomograms in predicting one-, three-, and five-year CSS, respectively. OS, overall survival; CSS, colon-specific survival.

**Supplementary Figure 8.** Decision curve analysis for overall survival nomograms in the training cohort. A, patients at baseline; B, Patients who have survived for one year; C, Patients who have survived for three years; D, Patients who have survived for five years.

**Supplementary Figure 9.** Decision curve analysis for overall survival nomograms in the test cohort. A, patients at baseline; B, patients who have survived for one year; C, patients who have survived for three years; D, patients who have survived for five years.

**Supplementary Figure 10.** Decision curve analysis for colon-specific survival nomograms in the training cohort. A, patients at baseline; B, patients who have survived for one year; C, patients who have survived for three years; D, patients who have survived for five years.

**Supplementary Figure 11.** Decision curve analysis for colon-specific survival nomograms in the test cohort. A, patients at baseline; B, patients who have survived for one year; C, patients who have survived for three years; D, patients who have survived for five years.

